# An ICD-10-based algorithm for identifying the cause of acute respiratory failure in mechanically ventilated patients

**DOI:** 10.64898/2026.11.18.26368330

**Authors:** Jeremy M. Kahn, Colleen M. Casey, Janice Dsouza, Steven J. Martinez, Timothy D. Girard, Niall T. Prendergast, Billie S. Davis

## Abstract

**Rationale:** Acute respiratory failure requiring invasive mechanical ventilation is a common reason for admission to an intensive care unit. Epidemiological research is limited by the lack of scalable approaches for identifying the primary cause of acute respiratory failure in these patients.

**Objectives:** To develop and validate an algorithm for classifying mechanically ventilated patients by the primary cause of acute respiratory failure using International Classification of Diseases, Tenth Revision (ICD-10) diagnosis and procedure codes.

**Methods:** We identified adult patients undergoing invasive mechanical ventilation in the electronic health record of a multi-hospital health system in the United States. Using this cohort, we created a preliminary classification schema that assigned patients to one of eight mutually exclusive causes of acute respiratory failure (stroke, chronic lung disease, heart failure, trauma or overdose, pneumonia, non-pneumonia sepsis, post-operative, and other) based on the presence or absence of pre-specified ICD-10 codes. We compared the preliminary classifications to a clinical reference standard ascertained via manual chart review in a stratified random sample of the development cohort, refining the schema to optimize concordance. We then assessed content validity by examining clinical characteristics and outcomes across diagnosis categories, both in the development cohort and in an external all-payer claims dataset.

**Results:** Of 8,130 hospitalizations in the development sample, all were successfully categorized into eight mutually exclusive groups. Compared to the reference standard of manual chart review (n=206), overall agreement was moderate (kappa = 0.44, 95% confidence interval: 0.39–0.49). After refinement, overall agreement improved slightly (kappa = 0.46, 95% confidence interval: 0.41–0.52). The final positive predictive values ranged from 78.6% (pneumonia) to 32.3% (stroke). The largest diagnostic category was non-pneumonia sepsis (26.1%), and the smallest diagnostic category was heart failure (6.2%). Clinical characteristics and outcomes approximated those expected from prior literature and were similar in the external dataset.

**Conclusions:** A novel ICD-10-based algorithm for determining the primary cause of acute respiratory failure in patients receiving invasive mechanical ventilation was moderately accurate. This algorithm is potentially useful for facilitating large-scale epidemiological research in this population.

## INTRODUCTION

In the United States an estimated 1.6 million patients undergo invasive mechanical ventilation for acute respiratory failure in an intensive care unit (ICU) each year (1). Traditionally, our understanding of the epidemiology and outcomes of these patients has come from prospective observational studies and clinical trials examining the efficacy of specific treatments (2–6). Increasingly, however, researchers are using population-based administrative data, pragmatic clinical trials, and hybrid implementation-effectiveness trials to provide new epidemiological insights and generate robust evidence on novel treatment strategies (7–14).

These study designs have significantly expanded the scope and scale of critical care research. However, they share a limitation in that researchers currently have no practical way to report data on the inciting cause of acute respiratory failure for the enrolled patients (e.g., pneumonia, heart failure, etc.). As a result, clinicians, health care administrators, and other researchers have little ability to understand what types of patients were included in the research and whether the results are transferable to outside populations. To address this methodological gap, we sought to develop and validate a scalable approach for identifying the primary cause of acute respiratory failure in mechanically ventilated ICU patients using widely available International Classification of Diseases, Tenth Revision (ICD-10) diagnosis and procedure codes.

## METHODS

### Study design and data

We performed a retrospective cohort study to develop a novel algorithm to classify ICU patients receiving invasive mechanical ventilation for acute respiratory failure by their primary inciting cause. We used the electronic health record of the UPMC health system, a large multi-hospital health care provider in the mid-Atlantic region of the United States. We obtained the data from UPMC’s existing ICU clinical data warehouse, which contains detailed clinical and administrative data including vital signs, laboratory values, respiratory parameters, ventilator settings, and medications, as well as ICD-10 diagnosis and procedure codes assigned at the end of the hospitalization. The data underwent standard cleaning procedures and have been used extensively in prior research (15–17).

### Hospitals and patients

We included patients admitted during the calendar years 2023 and 2024 to 10 UPMC hospitals that used a single EHR (Cerner PowerChart). These hospitals represented a mix of academic and community hospitals in urban, suburban, and rural areas. We identified all adult patients (≥18 years of age) receiving invasive mechanical ventilation through an endotracheal tube using documentation from the respiratory therapy flowsheets, which includes both mode of respiratory support and the type of airway.

### Conceptual rationale for key design decisions

Our goal was to design an algorithm that assigned patients to a manageable number of mutually exclusive diagnostic categories in which each category reflected the most likely primary cause of acute respiratory failure during the patient’s hospitalization. This approach carries several inherent tradeoffs, which we acknowledge as potential limitations of this work.

First, some patients can have multiple causes of respiratory failure. For these patients, the process of identifying a primary cause among many causes is fundamentally subjective. We opted to identify only a single primary cause because that would be more useful in terms of characterizing cohorts compared to the alternative of reporting multiple causes of respiratory failure for each patient. In making this decision, we intentionally accepted the tradeoff that potentially valuable information about the etiology of respiratory failure may be obscured by only allowing each patient to have a single cause.

Second, patients can experience multiple episodes of mechanical ventilation over the course of a hospitalization, each with a potentially different. As a result, important information may be obscured by assigning a single cause for each hospitalization. However, ICD codes are only reported at the level of the hospitalization, not the episode of mechanical ventilation, making this decision a necessity for our study design. In making this decision, we intentionally accepted the tradeoff of potential misclassification for patients with multiple episodes of mechanical ventilation within a given hospitalization.

Third, the true number of potential causes of acute respiratory failure is greater than can easily be grouped into a modest number of categories. As a result, any manageable categorization schema will necessarily represent an oversimplification of each patient’s complex pathophysiology. We chose to assign patients to eight categories—seven diagnostic categories (stroke, chronic lung disease, heart failure, pneumonia, trauma or overdose, post-operative, and non-pneumonia sepsis) and one “other” category for unclassified patients. This decision was in part conceptual and in part pragmatic, since we could only include diagnoses that could accurately be identified using ICD codes (18). In making this decision, we intentionally accepted the tradeoff that fewer categories means some loss of information, yet a lower possibility of misclassification.

### Initial algorithm development

To serve as building blocks for the algorithm, we first created flags for the following diagnoses using validated ICD-10 diagnosis codes: stroke (19, 20), chronic lung disease (21), heart failure (22), trauma or overdose (23), pneumonia (24), and non-pneumonia infections (25), noting whether the codes appeared in the primary position or a secondary position. We also created a flag for acute respiratory failure in any position (26), and a flag for major surgery using ICD-10 procedure codes based on the United States Agency for Healthcare Research and Ǫuality Healthcare Cost and Utilization Project Clinical Classification Software (27). The complete code sets and flag assignments are shown in **eTable 1** and **eTable 2**.

Using clinical judgment as a guide, we next used these flags to create a preliminary hierarchical algorithm that assigned patients to one of the seven diagnostic categories based on which combination of flags appeared in the primary or secondary positions (**eTable 3**). To ensure mutually exclusive assignments, once a hospitalization was assigned to a category, it could not be assigned to a second category. If patients did not receive an assignment via the algorithm, then they were assigned to the “other” category. As a result of this step, all hospitalizations were successfully assigned to a single mutually exclusive preliminary category (**eTable 4**).

### Reference standard ascertainment

As our reference standard, we used the consensus primary cause of acute respiratory failure as determined by two expert clinicians with experience caring for patients receiving mechanical ventilation in the ICU. To select cases for review we randomly sampled hospitalizations from the development cohort, stratifying by each diagnosis group (**eTable 5** and **eTable 6**). Based on power calculations designed to result in reasonable confidence intervals around the estimates of kappa, we planned to sample at least 200 charts. Ultimately, we sampled 228 charts so that there would be a relatively equal number of charts in each category.

Two independent reviewers then manually reviewed each sampled chart to determine the single best primary cause of respiratory failure for the first episode of mechanical ventilation for each hospitalization. There were four reviewers in total, so each reviewer reviewed 114 charts. To minimize the chance of bias, the reviewers were blinded to the algorithm-assigned category. Prior to chart review, all reviewers attended two training sessions in which they were instructed in standardized definitions of each category and received feedback on 10 practice chart reviews.

Following chart review, we used an iterative review process to determine the consensus category for each patient. First, we excluded 22 patients who, on chart review, were found not to have undergone invasive mechanical ventilation for acute respiratory failure during the hospitalization, resulting in a final sample of 206 patients. Next, we compared the two raters’ assessments. For patients for whom the two reviewers agreed on the cause (n=138 of 206, 67%, kappa=0.61), we considered that to be the consensus cause. For patients for whom the two reviewers initially disagreed (n=68 of 206, 33%), we asked them to discuss the case and come to an agreed-upon consensus cause. If they were able to do so (n=62 of 68, 91%), we considered that to be the consensus cause. If they were unable to do so (n=6 of 68, 9%), a third reviewer adjudicated the results and assigned the final cause. The result of this process was a single clinical reference standard representing the most likely primary cause of respiratory failure for each patient in the stratified random sample.

### Preliminary algorithm assessment and refinement

We compared the algorithm results to the reference standard by using confusion matrices, diagnostic performance tables, and the kappa statistic (28, 29). Based on the preliminary results (**eTable 7**, **eTable 8**, and **eTable G**), we examined whether specific changes to the algorithm would improve performance. We identified and tested changes that had the potential to increase the prevalence of categories with a high false negative rate (by moving them up in the order) and/or decrease the prevalence of categories with a low positive predictive value (by moving them down in the order). Only one of these changes improved the performance of the algorithm based on kappa (**eTable 10**) and was retained in the final assignment algorithm (**eTable 11**).

### Final algorithm assessment

We used a structured framework to assess the validity of the final algorithm. First, we examined criterion validity by comparing the accuracy of our final categorization relative to the reference standard using the same metrics as above (28). Second, we examined content validity by assessing the prevalence and clinical characteristics of each diagnostic group (30). We theorized that a valid algorithm would yield groups with prevalences and clinical characteristics roughly similar to those in the reported literature (2). Third, we examined transferability by applying the algorithm to an external cohort of discharges undergoing mechanical ventilation identified via ICD-10 procedure codes, and comparing the results to those in the development cohort (31). For this step, we used 2022 state inpatient discharge data from Maryland obtained from the United States Agency for Healthcare Research and Ǫuality Healthcare Cost and Utilization Project (HCUP). We chose Maryland because it is geographically adjacent and demographically similar to Pennsylvania, and we used 2022 because it was the most recent year of data available at the time of the study.

### Data management and regulatory review

Initial data processing was performed using Microsoft SǪL. Subsequent data management and analysis were performed using Stata Version 19.0. Stata code for the final algorithm is publicly available with the preprint of this manuscript. All aspects of this research were reviewed and approved by the University of Pittsburgh institutional review board (#23010181, covering use of UPMC data; and #26060206, covering use of HCUP data).

## RESULTS

A total of 7,530 patients with 8,130 hospitalizations were used to develop and validate the algorithm. Patient demographic and clinical characteristics are shown in **Table 1**. The final categorization is shown in **Table 2**. The final algorithm assigned 8.2% of hospitalizations to stroke, 9.1% of hospitalizations to chronic lung disease, 6.2% of hospitalizations to heart failure, 9.4% of hospitalizations to pneumonia, 15.1% of hospitalizations to trauma or overdose, 23.2% of hospitalizations to post-operative, 26.1% of hospitalizations to non-pneumonia sepsis, and 2.7% of hospitalizations to the other category. These categorizations were roughly similar to past published estimates (**Table 2**). Notable exceptions were non-pneumonia sepsis, which was considerably more common in the development cohort relative to the two comparator cohorts, and the “other” category, which was considerably less common in the development cohort relative to the two comparator cohorts.

**Table 1.**
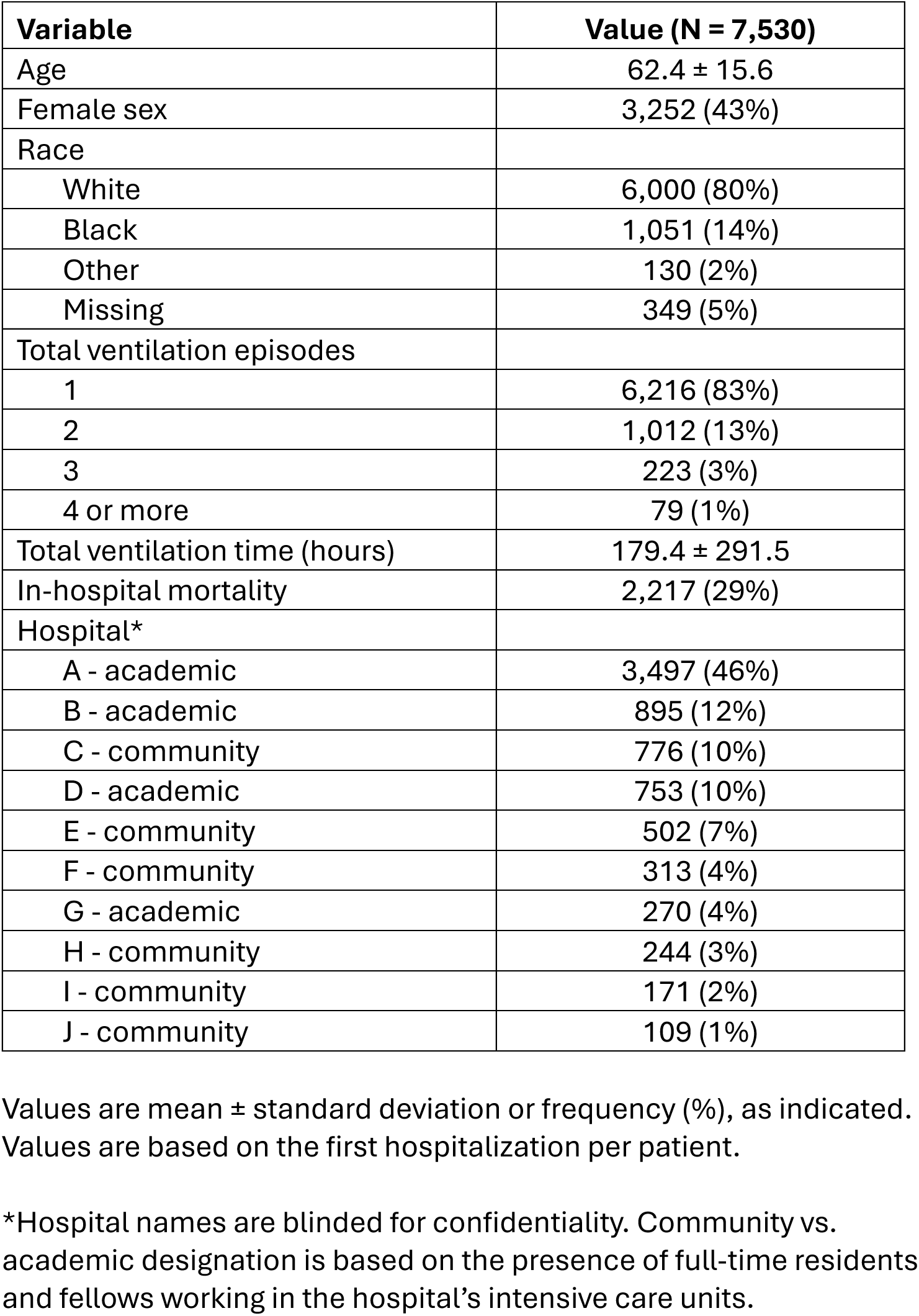
Characteristics of patients in the development cohort.

| **Variable** | **Value (N = 7,530)** |
|---|---|
| Age | 62.4 ± 15.6 |
| Female sex | 3,252 (43%) |
| Race |  |
| White | 6,000 (80%) |
| Black | 1,051 (14%) |
| Other | 130 (2%) |
| Missing | 349 (5%) |
| Total ventilation episodes |  |
| 1 | 6,216 (83%) |
| 2 | 1,012 (13%) |
| 3 | 223 (3%) |
| 4 or more | 79 (1%) |
| Total ventilation time (hours) | 179.4 ± 291.5 |
| In-hospital mortality | 2,217 (29%) |
| Hospital* |  |
| A - academic | 3,497 (46%) |
| B - academic | 895 (12%) |
| C - community | 776 (10%) |
| D - academic | 753 (10%) |
| E - community | 502 (7%) |
| F - community | 313 (4%) |
| G - academic | 270 (4%) |
| H - community | 244 (3%) |
| I - community | 171 (2%) |
| J - community | 109 (1%) |

Values are mean ± standard deviation or frequency (%), as indicated. Values are based on the first hospitalization per patient.
\*Hospital names are blinded for confidentiality. Community vs. academic designation is based on the presence of full-time residents and fellows working in the hospital’s intensive care units.

**Table 2.**
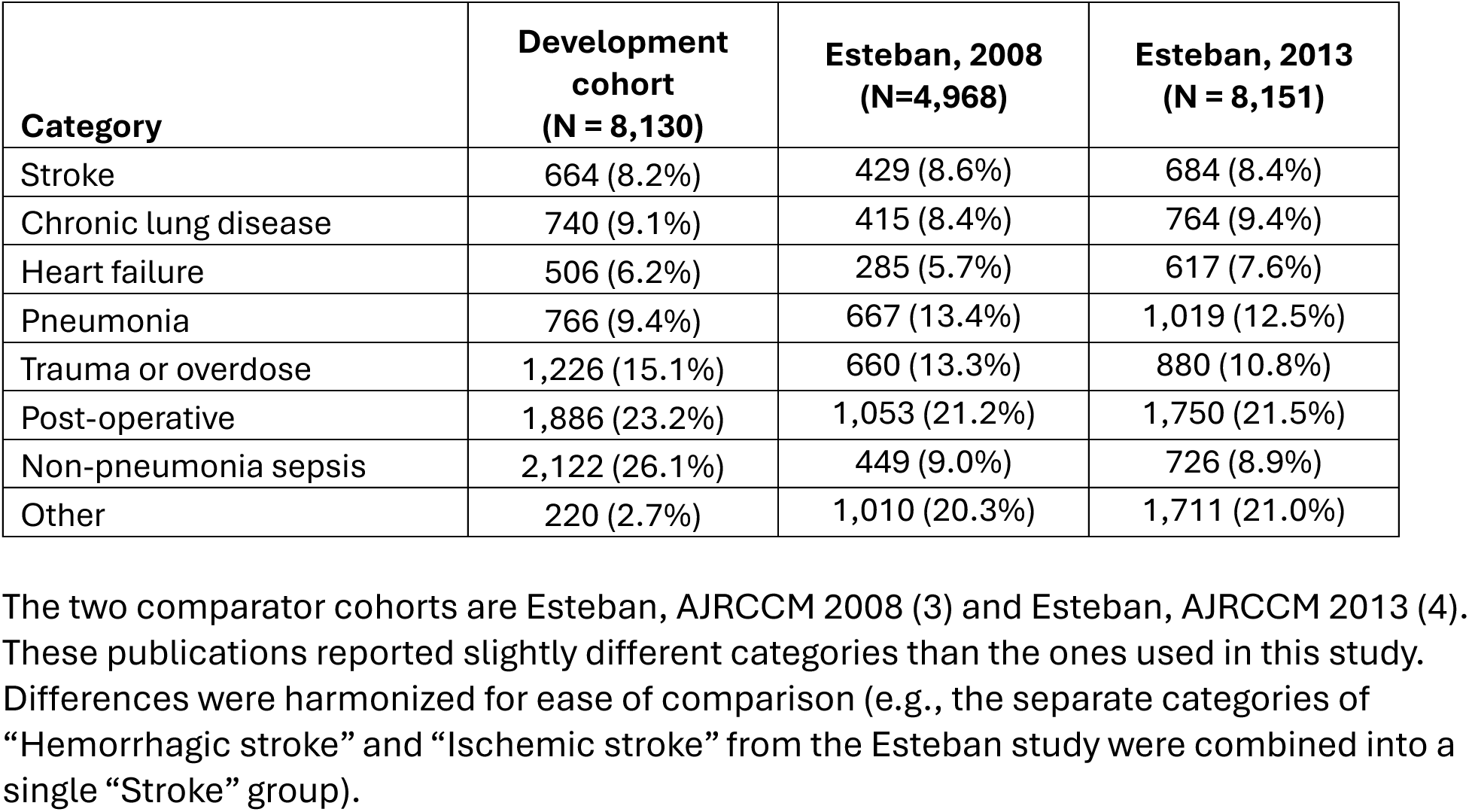
Diagnostic category assignments based upon the final hierarchical algorithm applied to the development cohort, alongside two comparator cohorts.

| **Category** | **Development cohort<br>(N = 8,130)** | **Esteban, 2008<br>(N=4,968)** | **Esteban, 2013<br>(N = 8,151)** |
|---|---|---|---|
| Stroke | 664 (8.2%) | 429 (8.6%) | 684 (8.4%) |
| Chronic lung disease | 740 (9.1%) | 415 (8.4%) | 764 (9.4%) |
| Heart failure | 506 (6.2%) | 285 (5.7%) | 617 (7.6%) |
| Pneumonia | 766 (9.4%) | 667 (13.4%) | 1,019 (12.5%) |
| Trauma or overdose | 1,226 (15.1%) | 660 (13.3%) | 880 (10.8%) |
| Post-operative | 1,886 (23.2%) | 1,053 (21.2%) | 1,750 (21.5%) |
| Non-pneumonia sepsis | 2,122 (26.1%) | 449 (9.0%) | 726 (8.9%) |
| Other | 220 (2.7%) | 1,010 (20.3%) | 1,711 (21.0%) |

The two comparator cohorts are Esteban, AJRCCM 2008 (3) and Esteban, AJRCCM 2013 (4). These publications reported slightly different categories than the ones used in this study. Differences were harmonized for ease of comparison (e.g., the separate categories of “Hemorrhagic stroke” and “Ischemic stroke” from the Esteban study were combined into a single “Stroke” group).

The confusion matrix and diagnostic performance table assessing the final categorization are shown in **Figure 1** and **Table 3**, respectively. False negative rates ranged from 0% (stroke) to 59.3% (pneumonia). Positive predictive values ranged from 78.6% (pneumonia) to 32.3% (stroke). Overall agreement was moderate (kappa = 0.46, 95% confidence interval: 0.41–0.52).

**Figure 1.**
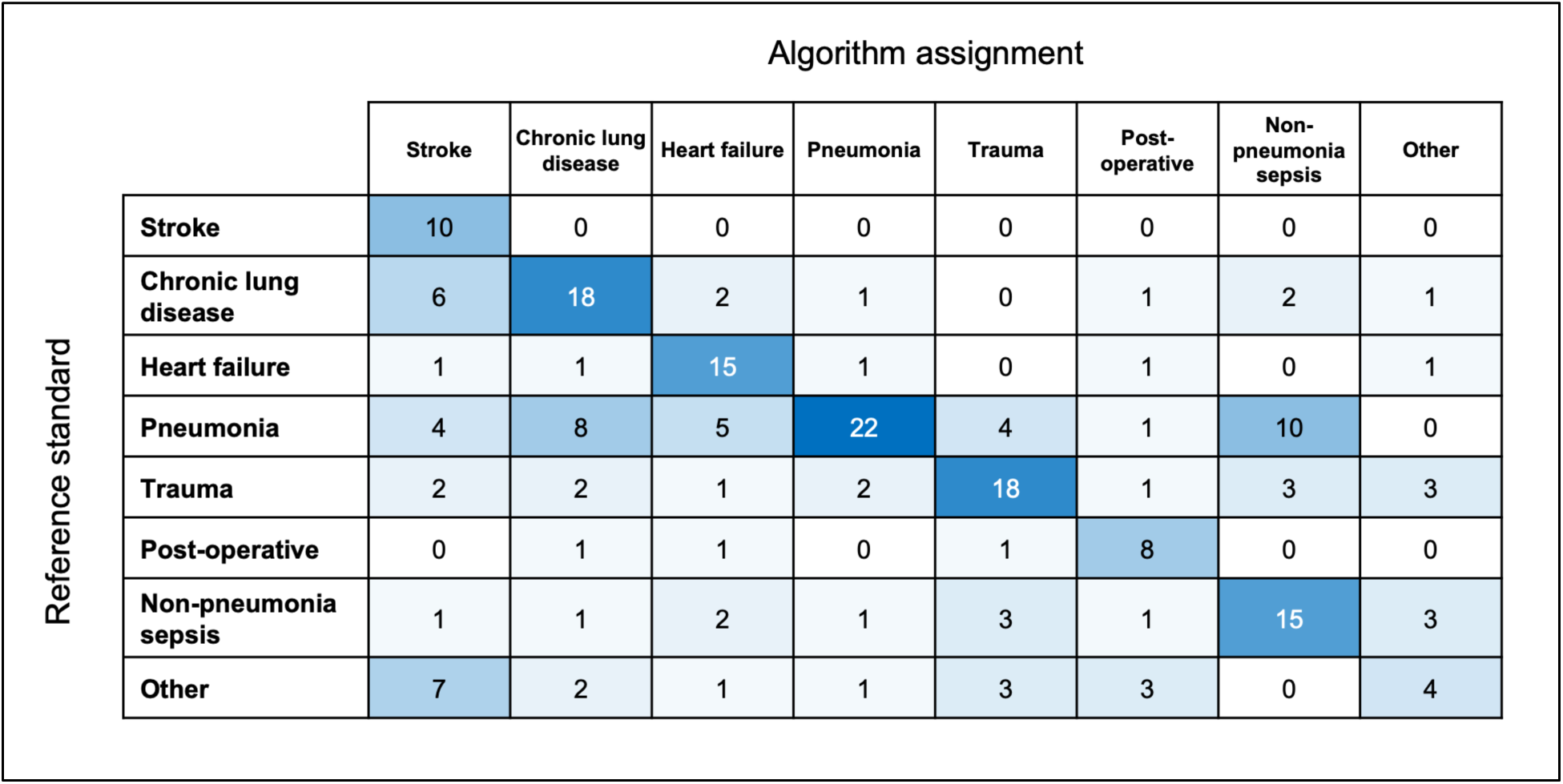
Confusion matrix and heatmap comparing the final algorithm-assigned diagnostic category (x-axis) with the reference standard diagnostic category (y-axis) in the development cohort

**Table 3.** Diagnostic performance for the final algorithm applied to the development cohort*.

|  | **Reference standard (N)** | **True positives (N)** | **Sensitivity (%)** | **False negative rate (%)** | **Positive predictive value (%)** |
|---|---|---|---|---|---|
| Stroke | 10 | 10 | 100 | 0 | 32.3 |
| CLD | 31 | 18 | 58.1 | 41.9 | 54.5 |
| Heart failure | 20 | 15 | 75.0 | 25.0 | 55.6 |
| Pneumonia | 54 | 22 | 40.7 | 59.3 | 78.6 |
| Trauma | 32 | 18 | 56.2 | 43.8 | 62.1 |
| Post-operative | 11 | 8 | 72.7 | 27.3 | 50.0 |
| NPS | 27 | 15 | 55.6 | 44.4 | 50.0 |
| Other | 21 | 4 | 19.0 | 81.0 | 33.3 |

CLD = chronic lung disease; NPS = non-pneumonia sepsis
\* Reference standard: the total number of patients based on chart review as the reference standard; true positives: the number of patients the algorithm correctly identified; sensitivity: the percentage of true cases caught by the algorithm; false negative rate: the percentage of true cases the algorithm missed; positive predictive value: the percentage of times the algorithm was correct when it issued that categorization.

The assessment of key clinical characteristics and outcomes by algorithm grouping is shown in **Table 4**. Consistent with a prior report (2), stroke patients, heart failure patients, and sepsis patients experienced the highest in-hospital mortality (45%, 36%, and 37%, respectively), while trauma/overdose patients and post-operative patients experienced the lowest mortality (23% and 20%, respectively). The application of the algorithm in the external sample yielded group prevalence estimates and mortality rates similar to those in the development cohort (**Table 5**).

**Table 4.**
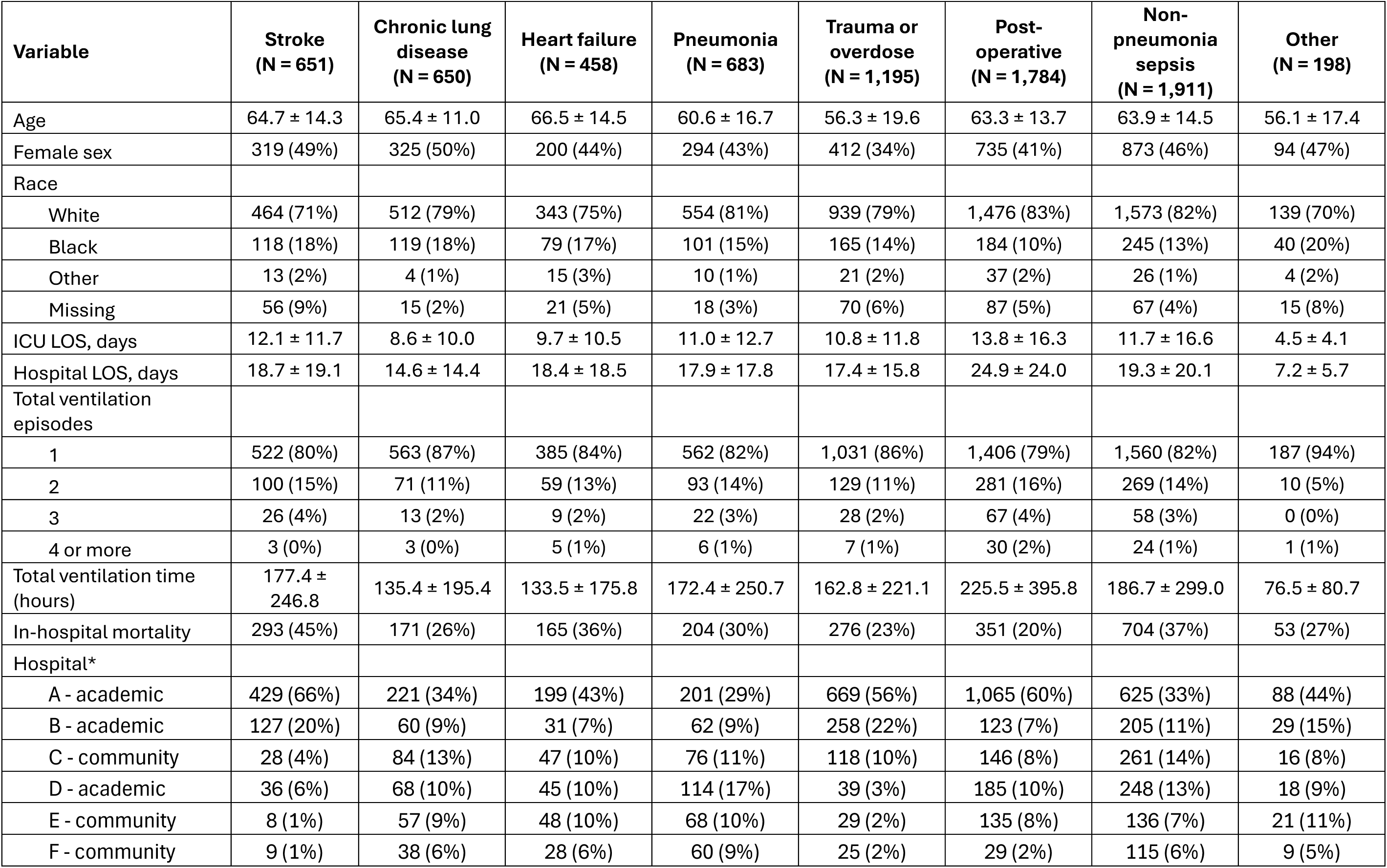
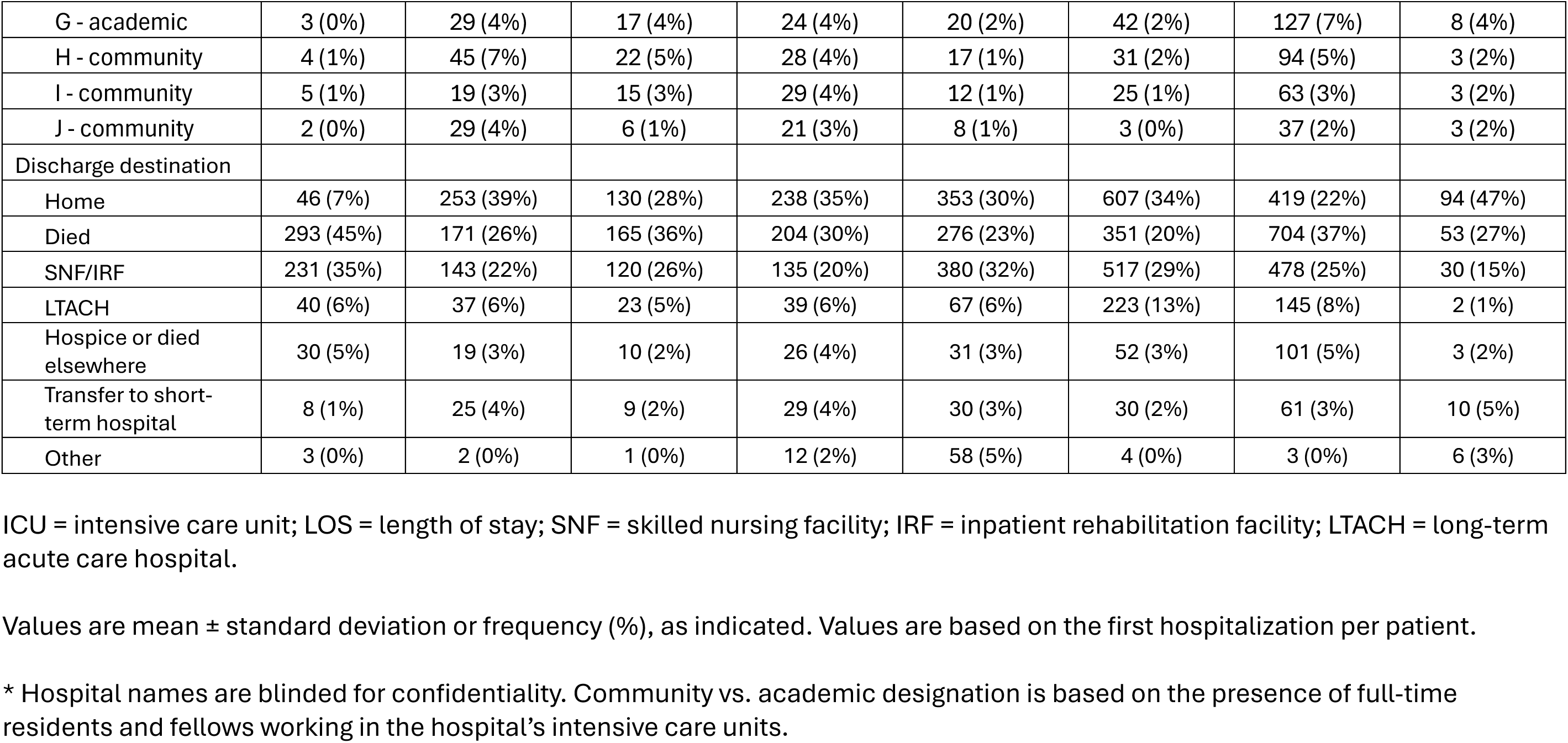
Patient characteristics by algorithm-assigned group in the development cohort.

| **Variable** | **Stroke<br>(N = 651)** | **Chronic lung disease<br>(N = 650)** | **Heart failure<br>(N = 458)** | **Pneumonia<br>(N = 683)** | **Trauma or overdose<br>(N = 1,195)** | **Post-operative<br>(N = 1,784)** | **Non-pneumonia sepsis<br>(N = 1,911)** | **Other<br>(N = 198)** |
|---|---|---|---|---|---|---|---|---|
| Age | 64.7 ± 14.3 | 65.4 ± 11.0 | 66.5 ± 14.5 | 60.6 ± 16.7 | 56.3 ± 19.6 | 63.3 ± 13.7 | 63.9 ± 14.5 | 56.1 ± 17.4 |
| Female sex | 319 (49%) | 325 (50%) | 200 (44%) | 294 (43%) | 412 (34%) | 735 (41%) | 873 (46%) | 94 (47%) |
| Race |  |  |  |  |  |  |  |  |
| White | 464 (71%) | 512 (79%) | 343 (75%) | 554 (81%) | 939 (79%) | 1,476 (83%) | 1,573 (82%) | 139 (70%) |
| Black | 118 (18%) | 119 (18%) | 79 (17%) | 101 (15%) | 165 (14%) | 184 (10%) | 245 (13%) | 40 (20%) |
| Other | 13 (2%) | 4 (1%) | 15 (3%) | 10 (1%) | 21 (2%) | 37 (2%) | 26 (1%) | 4 (2%) |
| Missing | 56 (9%) | 15 (2%) | 21 (5%) | 18 (3%) | 70 (6%) | 87 (5%) | 67 (4%) | 15 (8%) |
| ICU LOS, days | 12.1 ± 11.7 | 8.6 ± 10.0 | 9.7 ± 10.5 | 11.0 ± 12.7 | 10.8 ± 11.8 | 13.8 ± 16.3 | 11.7 ± 16.6 | 4.5 ± 4.1 |
| Hospital LOS, days | 18.7 ± 19.1 | 14.6 ± 14.4 | 18.4 ± 18.5 | 17.9 ± 17.8 | 17.4 ± 15.8 | 24.9 ± 24.0 | 19.3 ± 20.1 | 7.2 ± 5.7 |
| Total ventilation episodes |  |  |  |  |  |  |  |  |
| 1 | 522 (80%) | 563 (87%) | 385 (84%) | 562 (82%) | 1,031 (86%) | 1,406 (79%) | 1,560 (82%) | 187 (94%) |
| 2 | 100 (15%) | 71 (11%) | 59 (13%) | 93 (14%) | 129 (11%) | 281 (16%) | 269 (14%) | 10 (5%) |
| 3 | 26 (4%) | 13 (2%) | 9 (2%) | 22 (3%) | 28 (2%) | 67 (4%) | 58 (3%) | 0 (0%) |
| 4 or more | 3 (0%) | 3 (0%) | 5 (1%) | 6 (1%) | 7 (1%) | 30 (2%) | 24 (1%) | 1 (1%) |
| Total ventilation time (hours) | 177.4 ± 246.8 | 135.4 ± 195.4 | 133.5 ± 175.8 | 172.4 ± 250.7 | 162.8 ± 221.1 | 225.5 ± 395.8 | 186.7 ± 299.0 | 76.5 ± 80.7 |
| In-hospital mortality | 293 (45%) | 171 (26%) | 165 (36%) | 204 (30%) | 276 (23%) | 351 (20%) | 704 (37%) | 53 (27%) |
| Hospital* |  |  |  |  |  |  |  |  |
| A - academic | 429 (66%) | 221 (34%) | 199 (43%) | 201 (29%) | 669 (56%) | 1,065 (60%) | 625 (33%) | 88 (44%) |
| B - academic | 127 (20%) | 60 (9%) | 31 (7%) | 62 (9%) | 258 (22%) | 123 (7%) | 205 (11%) | 29 (15%) |
| C - community | 28 (4%) | 84 (13%) | 47 (10%) | 76 (11%) | 118 (10%) | 146 (8%) | 261 (14%) | 16 (8%) |
| D - academic | 36 (6%) | 68 (10%) | 45 (10%) | 114 (17%) | 39 (3%) | 185 (10%) | 248 (13%) | 18 (9%) |
| E - community | 8 (1%) | 57 (9%) | 48 (10%) | 68 (10%) | 29 (2%) | 135 (8%) | 136 (7%) | 21 (11%) |
| F - community | 9 (1%) | 38 (6%) | 28 (6%) | 60 (9%) | 25 (2%) | 29 (2%) | 115 (6%) | 9 (5%) |
| G - academic | 3 (0%) | 29 (4%) | 17 (4%) | 24 (4%) | 20 (2%) | 42 (2%) | 127 (7%) | 8 (4%) |
| H - community | 4 (1%) | 45 (7%) | 22 (5%) | 28 (4%) | 17 (1%) | 31 (2%) | 94 (5%) | 3 (2%) |
| I - community | 5 (1%) | 19 (3%) | 15 (3%) | 29 (4%) | 12 (1%) | 25 (1%) | 63 (3%) | 3 (2%) |
| J - community | 2 (0%) | 29 (4%) | 6 (1%) | 21 (3%) | 8 (1%) | 3 (0%) | 37 (2%) | 3 (2%) |
| Discharge destination |  |  |  |  |  |  |  |  |
| Home | 46 (7%) | 253 (39%) | 130 (28%) | 238 (35%) | 353 (30%) | 607 (34%) | 419 (22%) | 94 (47%) |
| Died | 293 (45%) | 171 (26%) | 165 (36%) | 204 (30%) | 276 (23%) | 351 (20%) | 704 (37%) | 53 (27%) |
| SNF/IRF | 231 (35%) | 143 (22%) | 120 (26%) | 135 (20%) | 380 (32%) | 517 (29%) | 478 (25%) | 30 (15%) |
| LTACH | 40 (6%) | 37 (6%) | 23 (5%) | 39 (6%) | 67 (6%) | 223 (13%) | 145 (8%) | 2 (1%) |
| Hospice or died elsewhere | 30 (5%) | 19 (3%) | 10 (2%) | 26 (4%) | 31 (3%) | 52 (3%) | 101 (5%) | 3 (2%) |
| Transfer to short-term hospital | 8 (1%) | 25 (4%) | 9 (2%) | 29 (4%) | 30 (3%) | 30 (2%) | 61 (3%) | 10 (5%) |
| Other | 3 (0%) | 2 (0%) | 1 (0%) | 12 (2%) | 58 (5%) | 4 (0%) | 3 (0%) | 6 (3%) |

ICU = intensive care unit; LOS = length of stay; SNF = skilled nursing facility; IRF = inpatient rehabilitation facility; LTACH = long-term acute care hospital.
Values are mean ± standard deviation or frequency (%), as indicated. Values are based on the first hospitalization per patient.
\* Hospital names are blinded for confidentiality. Community vs. academic designation is based on the presence of full-time residents and fellows working in the hospital’s intensive care units.

**Table 5.** Comparison between algorithm results in the development cohort and the external cohort*.

|  | **Development cohort** |  |  | **External cohort** |  |  |
|---|---|---|---|---|---|---|
| **Category** | **N** | **% of total** | **Mortality** | **N** | **% of total** | **Mortality** |
| Stroke | 651 | 9% | 45% | 1,226 | 8% | 38% |
| Chronic lung disease | 650 | 9% | 26% | 1,321 | 9% | 27% |
| Heart failure | 458 | 6% | 36% | 1,107 | 7% | 37% |
| Pneumonia | 683 | 9% | 30% | 1,888 | 12% | 40% |
| Trauma or overdose | 1,195 | 16% | 23% | 2,026 | 13% | 23% |
| Post-operative | 1,784 | 24% | 20% | 1,955 | 13% | 25% |
| Non-pneumonia sepsis | 1,911 | 25% | 37% | 5,189 | 34% | 41% |
| Other | 198 | 3% | 27% | 543 | 4% | 29% |

\* The external cohort is from the 2022 Maryland State Inpatient Database obtained from the Agency for Healthcare Research and Quality’s Healthcare Cost and Utilization Project. All values are for the first hospitalization per patient.

## DISCUSSION

We developed a novel algorithm for classifying mechanically ventilated patients by the primary cause of acute respiratory failure using International Classification of Diseases, Tenth Revision (ICD-10) diagnosis and procedure codes. The final algorithm classifies patients into eight mutually exclusive categories that have strong content validity and moderate concordance with categories determined via manual chart review.

This algorithm has several potential uses in epidemiological research, including characterizing cohorts in large, population-based studies using administrative data, pragmatic clinical trials using electronic health record data, and hybrid implementation-effectiveness studies conducted at the health system level. Better characterizing the patients in such studies will provide valuable context for the results and help readers understand if the study findings are transferable to different populations. In this respect, our algorithm fills a previously unmet need for ICU outcomes researchers.

That said, accuracy compared with our reference standard was only moderate, with a non-trivial amount of misclassification across all categories. Some misclassification is to be expected in any study due to the inherent limitations of ICD-10 codes, which are primarily used for billing purposes and therefore are an imperfect reflection of a patient’s clinical course. Additional known sources of misclassification include the fact that patients could have more than one primary cause of acute respiratory failure and more than one episode of mechanical ventilation during a hospitalization. Finally, there is always subjectivity inherent in diagnostic assessments, even those performed at the bedside, making it unlikely that any approach could achieve a high degree of accuracy.

Due to the risk of misclassification, we recommend that this algorithm be used primarily for the purposes of characterizing diverse cohorts of mechanically ventilated ICU patients, such as in the “Table 1” of a clinical trial or cohort study. When used in this way, the algorithm output should provide a reasonable reflection of the study’s population. This algorithm should not be used to identify specific types of patients for dedicated study, as the algorithm’s performance is probably insufficient for that purpose. In addition, this algorithm should not be used for risk adjustment, because the ICD-10 codes used are assigned at the end of the hospitalization and could plausibly reflect downstream consequences of exposures under study rather than variation in baseline risk.

Our study has several limitations. First, we developed the algorithm in a single health system, albeit one that is large, diverse in hospital type, and likely representative of most acute care settings in high-income countries. Future work should establish the performance of this algorithm in other settings, such as low-and middle-income countries. Second, we assessed the performance of our algorithm against an imperfect reference standard, albeit one that was obtained using rigorous and reproducible methodology.

Indeed, a consensus diagnosis among the first two experts could not be achieved in only six patients, or 2.9% of the total validation sample. Third, we made several design decisions that could have reduced the performance of our algorithm, including limiting each patient to only one diagnosis and omitting diagnoses that cannot be reliably assessed using ICD-10 codes, such as acute respiratory distress syndrome (32). However, these decisions were made in keeping with our overall goals for the algorithm and our intent to create a useful algorithm in which perfect did not become the enemy of the good.

## CONCLUSION

A novel ICD-10-based algorithm for classifying mechanically ventilated patients by the primary cause of acute respiratory failure demonstrated strong content validity and reasonable criterion validity. This algorithm will enable researchers to better describe their cohorts in large studies of mechanically ventilated patients, increasing the value of pragmatic outcomes research to the critical care community.

## Author contributions

All authors made substantial contributions to the conception and design of the work, the analysis and interpretation of the data. All authors critically revised the manuscript for important intellectual comment. JMK and BSD acquired the data. JMK wrote the initial manuscript.

## Disclosure of generative AI use

Claude Sonnet 5 (Anthropic) was used to proofread the manuscript for typographical and grammatical errors. There was no other use of generative AI in this work.

## Trial registration

not applicable

## Supporting information

eTable 1

eTable 2

eTable 3

eTable 4

eTable 5

eTable 6

eTable 7

eTable 8

eTable 9

eTable 10

eTable 11

Stata code for the final algorithm

## Data Availability

All data produced in the present study are available upon reasonable request to the authors, or are publicly available upon execution of a valid data use agreement with the data owners.

