## Supplementary material for "An ICD-10-based algorithm for identifying the cause of acute respiratory failure in mechanically ventilated patients": eTable 1

**eTable 1.** Complete set of ICD-10 codes used to create the initial flags

| **Flag** | **Codes (diagnoses codes unless otherwise specified)** * |
|---|---|
| Acute respiratory failure | J80, J96.00, J96.01, J96.02, J96.20, J96.21, J96.22, J96.90, J96.91, J96.92 |
| Heart failure | I11.0, I13.0, I13.2, 150* |
| Non-pneumonia infections | A* (excluding A48.1), B* (excluding B01.2, B05.2, B20.6), D73.3, G00*, G01, G02, I33.0, K35*, K36, K37, K55.3*, K57.00, K57.01, K57.12, K57.13, K57.20, K57.21, K57.32, K57.33, K57.40, K57.41, K57.52, K57.53, K57.80, K57.81, K57.92, K57.93, K61*, K63.0, K63.1, K65*, K67, K68.1*, K75.0, K81*, K85.02, K85.12, K85.22, K85.32, K85.82, K85.92, M72.6, N10, N30*, N34.0, O03.37, O03.38, O03.5, O03.87, O03.88, O04.87, O04.88, O07.37, O07.38, O08.82, O08.83, O23*, O41.1*, O85, O86*, O87.0, O87.1, O88.3*, O91*, O98*, R65.20, R65.21, R78.81 |
| Chronic lung disease | J40, J41.0, J41.1, J41.8, J42, J43.0, J43.1, J43.2, J43.8, J43.9, J44.0, J44.1, J44.89, J44.9, J45.21, J45.22, J45.31, J45.32, J45.41, J45.42, J45.51, J45.52, J45.901, J45.902, J45.990, J47.0, J47.1 |
| Pneumonia | A48.1, B01.2, B05.2, B20.6, J09.X1, J10.00, J10.01, J10.08, J11.00, J11.08, J12.0, J12.1, J12.2, J12.3, J12.81, J12.82, J12.89, J12.9, J13, J14, J15.0, J15.1, J15.20, J15.211, J15.212, J15.29, J15.3, J15.4, J15.5, J15.6, J15.7, J15.8, J15.9, J16.0, J16.8, J17, J17.0, J17.1, J17.2, J17.3, J17.8, J18, J18.0, J18.1, J18.8, J18.9, J69.0, J85.1, J86.0, J86.9, U07.1 |
| Stroke | I60*, I61*, I62*, I63* |
| Surgery | Major Procedure as mapped from Procedure Classes Refined for ICD-10-PCS (Procedure Coding System). Healthcare Cost and Utilization Project. January 2025. Agency for Healthcare Research and Quality, Rockville, MD. www.hcup-us.ahrq.gov/toolssoftware/procedureicd10/procedure_icd10.jsp; this software uses ICD-10 procedure codes, all positions |
| Trauma or overdose | S*, T* (excluding T8*), V*, W*, X*, Y00* to Y38* |

* An asterisk after a code indicates that all subcodes were included, with exceptions as noted.
