## Supplementary material for "An ICD-10-based algorithm for identifying the cause of acute respiratory failure in mechanically ventilated patients": eTable 2

**eTable 2.** Initial flag assignments in the development cohort (N=8,130).*

| Flag | Primary | Secondary |
|---|---|---|
| Acute respiratory failure | 520 (6%) | 6,218 (76%) |
| Heart failure | 189 (2%) | 2,836 (35%) |
| Non-pneumonia infections | 2,018 (25%) | 4,474 (55%) |
| Chronic lung disease | 77 (1%) | 2,554 (31%) |
| Pneumonia | 394 (5%) | 4,059 (50%) |
| Stroke | 554 (7%) | 829 (10%) |
| Surgery** | 3,822 (47%) | |
| Trauma or overdose | 1,077 (13%) | 2,793 (34%) |

* Flags are not mutually exclusive, either within patients (i.e., a patient could be flagged for multiple diagnoses) or across primary and secondary codes (i.e., a patient could be flagged for a given disease in both the primary position and the secondary position)

** Since the surgery flag is derived from procedure codes in any position, there are no primary and secondary designations.
