## Supplementary material for "An ICD-10-based algorithm for identifying the cause of acute respiratory failure in mechanically ventilated patients": eTable 3

**eTable 3.** Preliminary hierarchical algorithm applied to the development cohort (N=8,130) *

| Step | Primary | Secondary | Major Surgery | Preliminary Assignment | N (%) assigned | N (%) remaining |
|---|---|---|---|---|---|---|
| 1 | Stroke | | | Stroke | 554 (7%) | 7,576 (93%) |
| 2 | CLD | | | CLD | 77 (1%) | 7,499 (92%) |
| 3 | Heart failure | | | Heart failure | 189 (2%) | 7,310 (90%) |
| 4 | Pneumonia | | | Pneumonia | 394 (5%) | 6,916 (85%) |
| 5 | TOD | | | Trauma | 1,077 (13%) | 5,839 (72%) |
| 6 | ARF | | Yes | Post-operative | 77 (1%) | 5,762 (71%) |
| 7 | ARF | Stroke | | Stroke | 19 (<1%) | 5,743 (71%) |
| 8 | ARF | CLD | | CLD | 272 (3%) | 5,471 (67%) |
| 9 | ARF | Heart failure | | Heart failure | 57 (1%) | 5,414 (67%) |
| 10 | ARF | Pneumonia | | Pneumonia | 59 (1%) | 5,355 (66%) |
| 11 | ARF | TOD | | Trauma | 13 (<1%) | 5,342 (66%) |
| 12 | NPI | | | NPS | 2,018 (25%) | 3,324 (41%) |
| 13 | None | | Yes | Post-operative | 1,882 (23%) | 1,442 (18%) |
| 14 | None | Stroke | | Stroke | 86 (1%) | 1,356 (17%) |
| 15 | None | CLD | | CLD | 355 (4%) | 1,001 (12%) |
| 16 | None | Heart failure | | Heart failure | 252 (3%) | 749 (9%) |
| 17 | None | Pneumonia | | Pneumonia | 294 (4%) | 455 (6%) |
| 18 | None | TOD | | Trauma | 131 (2%) | 324 (4%) |
| 19 | None | NPI | | NPS | 104 (1%) | 220 (3%) |
| 20 | ARF or none | | | Other | 220 (3%) | 0 (0%) |

CLD = chronic lung disease; TOD = trauma or overdose; ARF = acute respiratory failure; NPI = non-pneumonia infection; NPS = non-pneumonia sepsis

* The preliminary algorithm was developed via the clinical judgement of the authors. Mutually exclusive diagnoses were assigned in order from 1 to 20. Columns provide the number and percentage of patients assigned to that diagnosis via each step, and the number and percentage of patients remaining in the pool after each step.
