## Supplementary material for "An ICD-10-based algorithm for identifying the cause of acute respiratory failure in mechanically ventilated patients": eTable 4

**eTable 4.** Preliminary categorization in the development cohort (N=8,130)*

| Preliminary category | N | % of total |
|---|---|---|
| Stroke | 659 | 8% |
| Chronic lung disease | 704 | 9% |
| Heart failure | 498 | 6% |
| Pneumonia | 747 | 9% |
| Trauma | 1,221 | 15% |
| Post-operative | 1,959 | 24% |
| Non-pneumonia sepsis | 2,122 | 26% |
| Other | 220 | 3% |

* This table shows the results of the preliminary assignments after applying the algorithm shown in eTable 3. These categories are mutually exclusive.
