## Supplementary material for "An ICD-10-based algorithm for identifying the cause of acute respiratory failure in mechanically ventilated patients": eTable 5

**eTable 5.** Sampling strategy by hierarchical step*

| Step | Primary | Secondary | Major Surgery | Preliminary Assignment | Charts sampled (N) |
|---|---|---|---|---|---|
| 1 | Stroke | | | Stroke | 10 |
| 2 | CLD | | | CLD | 10 |
| 3 | Heart failure | | | Heart failure | 10 |
| 4 | Pneumonia | | | Pneumonia | 10 |
| 5 | TOD | | | Trauma | 10 |
| 6 | ARF | | Yes | Post-operative | 15 |
| 7 | ARF | Stroke | | Stroke | 10 |
| 8 | ARF | CLD | | CLD | 10 |
| 9 | ARF | Heart failure | | Heart failure | 10 |
| 10 | ARF | Pneumonia | | Pneumonia | 10 |
| 11 | ARF | TOD | | Trauma | 10 |
| 12 | NPI | | | NPS | 15 |
| 13 | None | | Yes | Post-operative | 15 |
| 14 | None | Stroke | | Stroke | 10 |
| 15 | None | CLD | | CLD | 10 |
| 16 | None | Heart failure | | Heart failure | 10 |
| 17 | None | Pneumonia | | Pneumonia | 10 |
| 18 | None | TOD | | Trauma | 10 |
| 19 | None | NPI | | NPS | 15 |
| 20 | ARF or none | | | Other | 18 |

CLD = chronic lung disease; TOD = trauma or overdose; ARF = acute respiratory failure; NPI = non-pneumonia infection; NPS = non-pneumonia sepsis

* To obtain the sample for the reference standard ascertainment, we randomly sampled either 10, 15, or 18 charts from those assigned in each step in order to both evenly distribute the number of charts and reach our target sample size.
