## Supplementary material for "An ICD-10-based algorithm for identifying the cause of acute respiratory failure in mechanically ventilated patients": eTable 6

**eTable 6.** Sampling strategy by diagnostic category.

| Preliminary category | Hierarchy rows | Charts sampled (N) |
|---|---|---|
| Stroke | 3 | 30 (3 x 10) |
| Chronic lung disease | 3 | 30 (3 x 10) |
| Heart failure | 3 | 30 (3 x 10) |
| Pneumonia | 3 | 30 (3 x 10) |
| Trauma | 3 | 30 (3 x 10) |
| Post-operative | 2 | 30 (2 x 15) |
| Non-pneumonia sepsis | 2 | 30 (2 x 15) |
| Other | 1 | 18 |

* To obtain the sample for the reference standard ascertainment, we randomly sampled either 30 or 18 charts from each diagnosis in order to both evenly distribute the number of charts and reach our target sample size.
