## Supplementary material for "An ICD-10-based algorithm for identifying the cause of acute respiratory failure in mechanically ventilated patients": eTable 7

**eTable 7.** Confusion matrix and heatmap comparing the preliminary algorithm-assigned diagnostic category (x-axis) with the reference standard diagnostic category (y-axis) in the development cohort

| | **Algorithm assignment** | | | | | | | |
|---|---|---|---|---|---|---|---|---|
| **Reference standard** | Stroke | CLD | Heart failure | Pneu-monia | Trauma | Post-operative | NPS | Other |
| Stroke | 10 | 0 | 0 | 0 | 0 | 0 | 0 | 0 |
| CLD | 6 | 15 | 2 | 1 | 0 | 4 | 2 | 1 |
| Heart failure | 1 | 1 | 15 | 1 | 0 | 1 | 0 | 1 |
| Pneumonia | 4 | 7 | 4 | 19 | 4 | 6 | 10 | 0 |
| Trauma | 2 | 2 | 1 | 2 | 18 | 1 | 3 | 3 |
| Post-operative | 0 | 0 | 1 | 0 | 1 | 9 | 0 | 0 |
| NPS | 1 | 1 | 1 | 1 | 3 | 2 | 15 | 3 |
| Other | 6 | 1 | 1 | 1 | 3 | 5 | 0 | 4 |

CLD = chronic lung disease; NPS = non-pneumonia sepsis
