## Supplementary material for "An ICD-10-based algorithm for identifying the cause of acute respiratory failure in mechanically ventilated patients": eTable 8

**eTable 8.** Diagnostic performance table for the preliminary algorithm applied to the development cohort*

| | Reference standard (N) | True positives (N) | Sensitivity (%) | False negative rate (%) | Positive predictive value (%) |
|---|---|---|---|---|---|
| Stroke | 10 | 10 | 100 | 0 | 33.3 |
| CLD | 31 | 15 | 48.4 | 51.6 | 55.6 |
| Heart failure | 20 | 15 | 75.0 | 25.0 | 60.0 |
| Pneumonia | 54 | 19 | 35.2 | 64.8 | 76.0 |
| Trauma | 32 | 18 | 56.2 | 43.8 | 62.1 |
| Post-operative | 11 | 9 | 81.8 | 18.2 | 32.1 |
| NPS | 27 | 15 | 55.6 | 44.4 | 50.0 |
| Other | 21 | 4 | 19.0 | 81.0 | 33.3 |

CLD = chronic lung disease; NPS = non-pneumonia sepsis

* Reference standard: the total number of patients based on chart review as the reference standard; true positives: the number of patients the algorithm correctly identified; sensitivity: the percentage of true cases caught by the algorithm; false negative rate: the percentage of true cases the algorithm missed; positive predictive value: the percentage of times the algorithm was correct when it issued that categorization
