## Supplementary material for "An ICD-10-based algorithm for identifying the cause of acute respiratory failure in mechanically ventilated patients": eTable 9

**eTable 9.** Reliability metrics for the preliminary algorithm in the development cohort

| | Overall Accuracy | Baseline chance agreement | Kappa (95% CI) | p-value |
|---|---|---|---|---|
| Preliminary | 51.0% | 12.5% | 0.44 (0.39, 0.49) | p < 0.001 |

CI = confidence interval
