## Supplementary material for "An ICD-10-based algorithm for identifying the cause of acute respiratory failure in mechanically ventilated patients": eTable 10

**eTable 10.** Reliability metrics given potential changes to the algorithm in the development cohort.*

| | Overall Accuracy | Baseline chance agreement | Kappa (95% CI) | p-value |
|---|---|---|---|---|
| Preliminary | 51.0% | 12.5% | 0.44 (0.39, 0.49) | p < 0.001 |
| Change: A | 53.4% | 13.1% | 0.46 (0.41, 0.52) | p < 0.001 |
| Change: B | 49.0% | 13.4% | 0.41 (0.36, 0.46) | p < 0.001 |
| Change: C | 49.0% | 13.1% | 0.41 (0.36, 0.47) | p < 0.001 |
| Change: D | 51.0% | 14.6% | 0.43 (0.37, 0.48) | p < 0.001 |
| Change: A, B | 51.0% | 14.5% | 0.43 (0.37, 0.48) | p < 0.001 |
| Change: A, C | 51.5% | 13.7% | 0.44 (0.38, 0.49) | p < 0.001 |
| Change: A, D | 51.0% | 16.6% | 0.41 (0.36, 0.47) | p < 0.001 |
| Change: B, C | 47.1% | 14.0% | 0.38 (0.33, 0.44) | p < 0.001 |
| Change: B, D | 49.0% | 15.5% | 0.40 (0.34, 0.45) | p < 0.001 |
| Change: C, D | 47.6% | 15.7% | 0.38 (0.32, 0.43) | p < 0.001 |
| Change: A, B, C | 49.0% | 15.1% | 0.40 (0.34, 0.45) | p < 0.001 |
| Change: A, B, D | 51.0% | 16.6% | 0.41 (0.36, 0.47) | p < 0.001 |
| Change: A, C, D | 49.5% | 16.8% | 0.39 (0.34, 0.45) | p < 0.001 |
| Change: B, C, D | 47.6% | 15.7% | 0.38 (0.32, 0.43) | p < 0.001 |
| Change: A, B, C, D | 49.5% | 16.8% | 0.39 (0.34, 0.45) | p < 0.001 |

CI = confidence interval.

* Please see **eTable 3** for the preliminary algorithm. Change A reorders the steps so that step 6 is immediately above step 13; Change B reorders the steps so that step 10 is immediately above step 8; Change C reorders the steps so that step 13 is immediately above 20; Change D reorders the steps so that step 17 is immediately above step 8. These changes were selected based on their potential to increase the prevalence of categories with a high false negative rate (by moving them up in the order) and/or decrease the prevalence of categories with a low positive predictive value (by moving them down in the order). The table shows reliability metrics after each change, both in isolation and together with the other changes. Only Change A (shaded in blue) increased the kappa score relative to the preliminary algorithm. This change was retained for the final algorithm, which is shown in **eTable 11**.
