## Supplementary material for "An ICD-10-based algorithm for identifying the cause of acute respiratory failure in mechanically ventilated patients": eTable 11

**eTable 11.** Final hierarchical algorithm applied to the development cohort

| Step | Primary | Secondary | Major Surgery | Preliminary Assignment | N (%) assigned | N (%) remaining |
|---|---|---|---|---|---|---|
| 1 | Stroke | | | Stroke | 554 (7%) | 7,576 (93%) |
| 2 | CLD | | | CLD | 77 (1%) | 7,499 (92%) |
| 3 | Heart failure | | | Heart failure | 189 (2%) | 7,310 (90%) |
| 4 | Pneumonia | | | Pneumonia | 394 (5%) | 6,916 (85%) |
| 5 | TOD | | | Trauma | 1,077 (13%) | 5,839 (72%) |
| 6 | ARF | Stroke | | Stroke | 24 (0%) | 5,815 (72%) |
| 7 | ARF | CLD | | CLD | 308 4(%) | 5,507 (68%) |
| 8 | ARF | Heart failure | | Heart failure | 65 (1%) | 5,442 (67%) |
| 9 | ARF | Pneumonia | | Pneumonia | 78 (1%) | 5,364 (66%) |
| 10 | ARF | Trauma | | Trauma | 18 (<1%) | 5,346 (66%) |
| 11 | NPI | | | NPS | 2,018 (25%) | 3,328 (41%) |
| 12 | ARF | | Yes | Post-operative | 4 (<1%) | 3,324 (41%) |
| 13 | None | | Yes | Post-operative | 1,882 (23%) | 1,442 (18%) |
| 14 | None | Stroke | | Stroke | 86 (1%) | 1,356 (17%) |
| 15 | None | CLD | | CLD | 355 (4%) | 1,001 (12%) |
| 16 | None | Heart failure | | Heart failure | 252 (3%) | 749 (9%) |
| 17 | None | Pneumonia | | Pneumonia | 294 (4%) | 455 (6%) |
| 18 | None | TOD | | Trauma | 131 (2%) | 324 (4%) |
| 19 | None | NPI | | Non-pneumonia sepsis | 104 (1%) | 220 (3%) |
| 20 | ARF or none | | | Other | 220 (3%) | 0 (0%) |

CLD = chronic lung disease; TOD = trauma or overdose; ARF = acute respiratory failure; NPI = non-pneumonia infection; NPS = non-pneumonia sepsis

* The final algorithm shown here differs from the preliminary version in that line 12 here was previously line 6. Moving this line down in the algorithm results in fewer patients being assigned to the post-operative category in favor of higher categories.
